# Determinants of Post-acute COVID-19 Syndrome among hospitalized severe COVID-19 patients: a 2-year follow up study

**DOI:** 10.1101/2023.06.13.23290674

**Authors:** Tamrat P. Elias, Tsegaye W. Gebreamlak, Tigist T. Gebremeskel, Binyam L. Adde, Bitaniya P. Elias, Abel M. Argaw, Addis A. Tenaw

## Abstract

**Background:** Coronavirus disease-19 (COVID-19), emerged as a public health threat in December 2019. The number of COVID-19 cases worldwide is now more than 765 million with more than 6.9 million dead. During follow-up visits following discharge, a large percentage of patients were discovered to still be suffering from health issues that lower their quality of life and ability to return to work. This study assessed the prevalence and associated risk factors of post-acute COVID-19 syndrome (PACS) among severe COVID-19 patients who were discharged from Millennium COVID-19 care center, Addis Ababa, Ethiopia.

**Methods:** A cross-sectional study using data collected from patient charts and a follow-up telephone interview after two years of discharge. Systematic random samplingwas used to select a total of 400 patients. A structured questionnaire developed from the case report form for PACS of WHO was used. Frequency and cross-tabulation were used for descriptive statistics. Predictor variables with a p-value <0.25 in bivariate analyses were included in the logistic regression.

**Result:** Out of the 400 patients, 20 patients were dead, 14 patients refused to give consent, and 26 patients couldn’t be reached because their phone wasn’t working. Finally, 340 were included in the study. The majority (68.5%) were males and the mean age was 53.9 (±13.3 SD) years. Most of the patients (60%) has one or more comorbidity. The most common symptom at presentation was cough (93.5%), followed by shortness of breath (82.1%) and fatigue (69.7%). The mean duration of hospital admission was 12.3 (±6.5 SD) days. More than a third (38.1%) of the patients reported the persistence of at least one symptom after hospital discharge. The most common symptoms were fatigue (27.5%) and Cough (15.3%). older age (AOR 1.04, 95% CI 1.02 – 1.07), female sex (AOR 1.82, 95% CI 1.00 – 3.29), presence of comorbidity (AOR 2.38, 95% CI 1.35 – 4.19), alcohol use (AOR 3.05, 95% CI 1.49 – 6.26), fatigue at presentation (AOR 2.18, 95% CI 1.21 – 3.95), and longer hospital stay (AOR 1.06, 95% CI 1.02 – 1.10) were found to increase the odds of developing post-acute COVID-19 syndrome. Higher hemoglobin level was found to decrease the risk of subsequent post-acute COVID-19 syndrome (AOR 0.84, 95% CI 0.71 – 0.99).

**Conclusion:** establishing a dedicated PACS follow-up clinic, especially for those with a higher risk can help to provide comprehensive care for the patients and improve their quality of life.

## Introduction

### Background

SARS-CoV-2, which causes coronavirus disease-19 (COVID-19), emerged as a public health threat in December, 2019 (1). According to the online world health organization (WHO) COVID-19 dashboard, as of May 1, 2023, COVID-19 pandemic affected more than 765 million peoples and caused more than 6.9 million deaths globally (2). The detection and treatment of acute illness does not appear to be the end of the COVID-19 fight (3). It has lately come to light that some patients’ incapacitating symptoms might last for weeks or even months (4). This manifestation was termed ‘post-acute COVID-19 syndrome’, ‘post COVID conditions’, ‘chronic COVID-19’or ‘long COVID’ (5). The number of post-acute COVID-19 patients is rapidly increasing due to the fact that millions of people have already contracted the disease and that many more will do so in the future (6). The capacity of people to return to work can be seriously impacted by persistent COVID symptoms, with substantial psychological, social, and economic repercussions for those affected, their families, and society as a whole (7).

The prevalence and clinical presentation of PACS are highly heterogeneous. The most frequently reported symptoms are fatigue, cardio-respiratory problems, and neurological symptoms (8). There is a wide difference among the prevalence of post-acute COVID-19, from 46% in Bangladesh to 81% in Italy (9, 10). Some researchers concluded that female gender and older age are important risk factors for eventual PACS (11, 12), but others, found no link between these sociodemographic characteristics and the development of PACS (13). There is a significant difference in the literatures whether or not risk factors for developing PACS include the existence of comorbidities (11, 14), the type of symptoms that present during an acute illness (13, 15), the length of hospitalization (9, 13), and the amount of oxygen needed upon admission (16, 17). Cigarette smoking was not associated with PACS in some studies (18), while others found out a strong association (16, 19).

### Rationale

Ethiopia notified the first confirmed case of COVID-19 on March 13, 2020 (20). The country recorded the largest number of COVID-19 confirmed cases in East Africa (2), implying a large number of patients with PACS. There are no post-COVID clinics in Ethiopia, nor is there a documented guideline for the management of post-COVID sequelae. There were no research articles in peer-reviewed journals measuring the burden of PACS in Ethiopia at the time this study was conceived. To alleviate these issues, we need to understand the prevalence and risk factors of PACS in order to establish effective management measures.

### Objective

As a result, the aim of this study was to assess the prevalence and associated risk factors of PACS among severe COVID-19 patients who were discharged alive from Millennium COVID-19 Care and treatment center, Addis Ababa, Ethiopia between July 1, 2020 and November 1, 2021.

## Methodology

### Study design and setting

A cross-sectional study design was used to assess the prevalence and associated risk factors of post-acute COVID-19 syndrome among severe COVID-19 patients who were discharged alive from Millennium COVID-19 care center, Addis Ababa, Ethiopia. Millennium COVID-19 Care Center (MCCC), was a makeshift hospital in Addis Ababa, the capital city of Ethiopia. The center was the biggest COVID-19 treatment facility in the country. It began giving service on June 2, 2020, and according to the center’s health management information system report (HMIS), as of November 1, 2021, a total of 6,760 patients were admitted and 5,580 patients were discharged alive.

### Study Participants

From adult patients (>18 years of age) who were admitted to MCCC with the diagnosis of Severe COVID-19 infection, confirmed by polymerase chain reaction (PCR) or rapid diagnostic test (RDT), those who were discharged alive between June 12,2020 and November 1, 2021 were the study population.

### Data collection tools and procedures

The socio-demographic profiles, past medical history including comorbidity, duration of symptoms before hospital admission, length of hospital stay, maximum amount of oxygen required during hospital stay, acute manifestations of COVID-19, and baseline laboratory investigations were extracted from patient charts. After taking verbal consent, a detailed telephonic interview was conducted with the study participants between February 1, 2023 and April 30, 2023 to record self-reported PACS symptoms and their characteristics, and self-assessment of current health status compared to the pre-COVID state. Data on COVID-19 vaccination history and current substance use were also collected during the telephone interview. The questionnaire was adapted from the W.H.O Global COVID-19 Clinical Platform Case Report Form (CRF) for Post COVID condition (Post COVID-19 CRF) (21). Data collectors were given a training prior to the data collection. Pre-testing of the questionnaire was also done. The collected data were entered into Epi-info software version 7 and then exported to Statistical Package for Social Sciences (SPSS) version 25 for cleaning and analysis. Individual who could not be contacted after two attempts were excluded.

### Sample size and statistical analysis

The sample size was determined for the prevalence by using single population proportion formula and for the sociodemographic, clinical, and behavioral risk factors by using double population proportion formula. Sample size calculated for the prevalence by using the single population proportion formula by considering p = 50%,as the prevalence of post-acute COVID syndrome is not known in Ethiopia, 95% confidence level (Zα/2 = 1.96), and a 5% margin of error yields the largest sample size. Since the source population is less than ten thousand (6760), a population correction formula was used and 10% non-response rate was added to yield a final sample size of 400. Systematic random sampling technique was used to select the study participants.

Frequency and cross-tabulation are used to summarize descriptive statistics of the data. Associations between predictor variables and outcomes of interest are estimated using both bivariate analysis and binary logistic regression. Predictor variables with a p-value <0.25 in bivariate analyses are reported and included in the logistic regression. For the Binary Logistic regression, 95% confidence interval for adjusted odds ratio (AOR) was calculated and variables with p-value ≤ 0.05 were considered as statistically associated with PACS.

### Ethical consideration

The study was conducted after obtaining ethical clearance from St. Paul’s Hospital Millennium Medical College Institutional Review Board. Verbal informed consent was taken from study participants during the telephone interview after explaining the purpose and objectives of the study. Confidentiality of individual patient information is maintained by using code numbers instead of other identifiers and the information gained from the chart and phone call is used only for research purposes.

## Results

Out of the 400 patients selected for the study, 20 patients died after hospital discharge, 14 patients refused to give consent, and 26 patients couldn’t be reached because their phone wasn’t working. The study included a total of 340 patients who were admitted to Millennium COVID-19 Care and treatment center with the diagnosis of severe COVID-19 pneumonia and discharged alive between June 12, 2020, and November 1, 2021. The mean duration from hospital discharge to the interview was 25.6 (± 4.8) months.

### Baseline sociodemographic and clinical characteristics

The majority (68.5%) of the study participants were male and the remaining 31.5% were females. The mean age at the time of admission to the center was 53.9 (±13.3 SD) years. The minimum age was 22 years and the maximum age was 85 years.

More than half of the patients (60%) has one or more comorbidity. The most common comorbidity among the patients was diabetes (35.6%), followed by hypertension (34.1%), chronic heart disease (6.5%), asthma or COPD (5.6%), dyslipidemia (3.8%), HIV (3.2%), Cancer (1.5%), CLD (1.5%), stroke (1.2%), and CKD (0.3%).

**Table 1:**
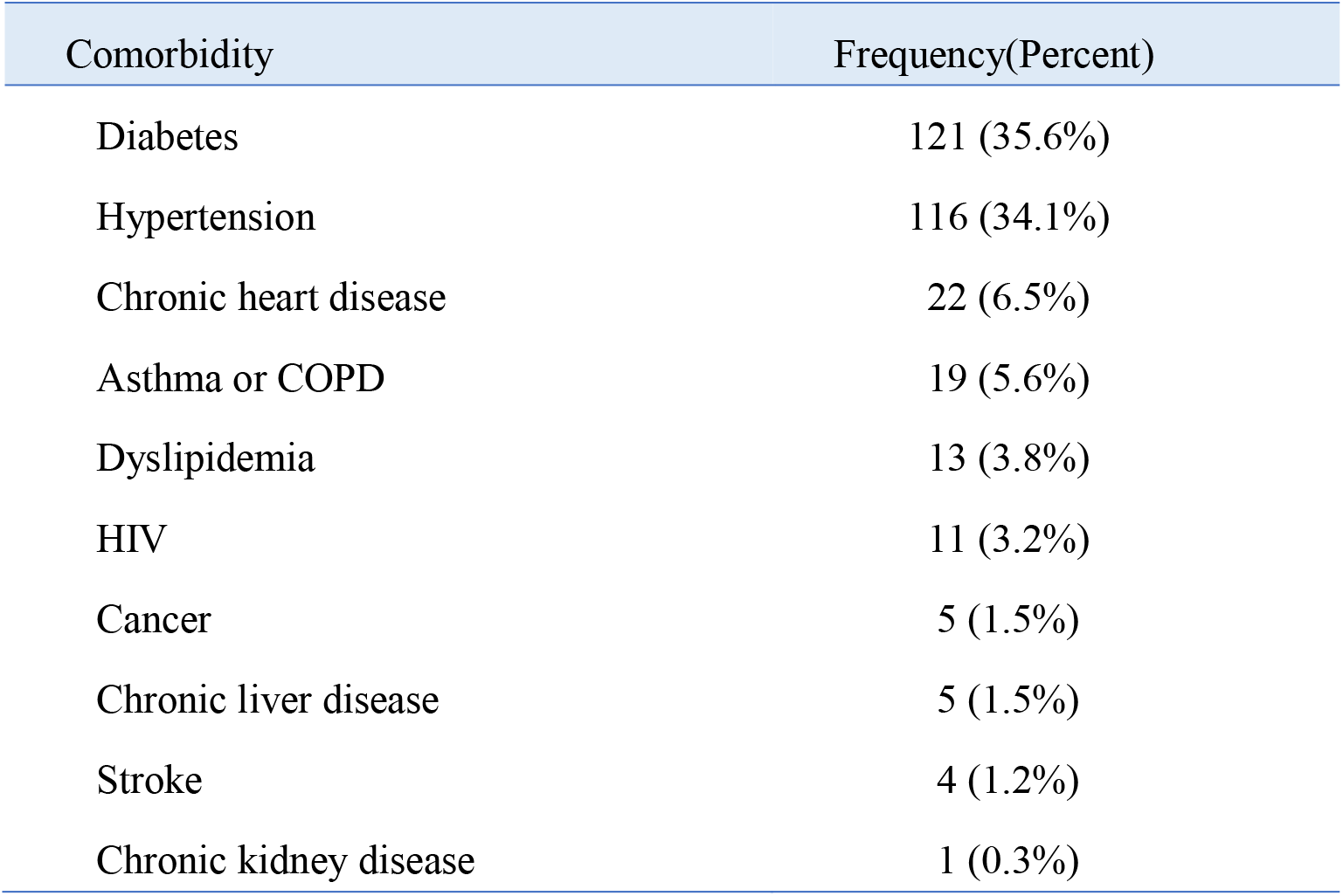
Comorbidity pattern of patients who were admitted at Millennium COVID-19 care and treatment center with the diagnosis of severe COVID-19 pneumonia (n=340).

| Comorbidity | Frequency(Percent) |
| --- | --- |
| Diabetes | 121 (35.6%) |
| Hypertension | 116 (34.1%) |
| Chronic heart disease | 22 (6.5%) |
| Asthma or COPD | 19 (5.6%) |
| Dyslipidemia | 13 (3.8%) |
| HIV | 11 (3.2%) |
| Cancer | 5 (1.5%) |
| Chronic liver disease | 5 (1.5%) |
| Stroke | 4 (1.2%) |
| Chronic kidney disease | 1 (0.3%) |

The mean duration of symptom onset before presentation was 8.3 (±4.1 SD) days. The minimum was 1 day and the maximum was 30 days. The mean duration of hospital admission was 12.3 (±6.5 SD) days. The minimum was 2 days and the maximum was 39 days. The mean maximum amount of oxygen required during a hospital stay was 5.8 (±4.0 SD) liters. The minimum oxygen requirement was 1 liter and the maximum was 15 liters.

The most common symptoms at presentation were cough (93.5%), followed by shortness of breath (82.1%), and fatigue (69.7%).

**Table 2:**
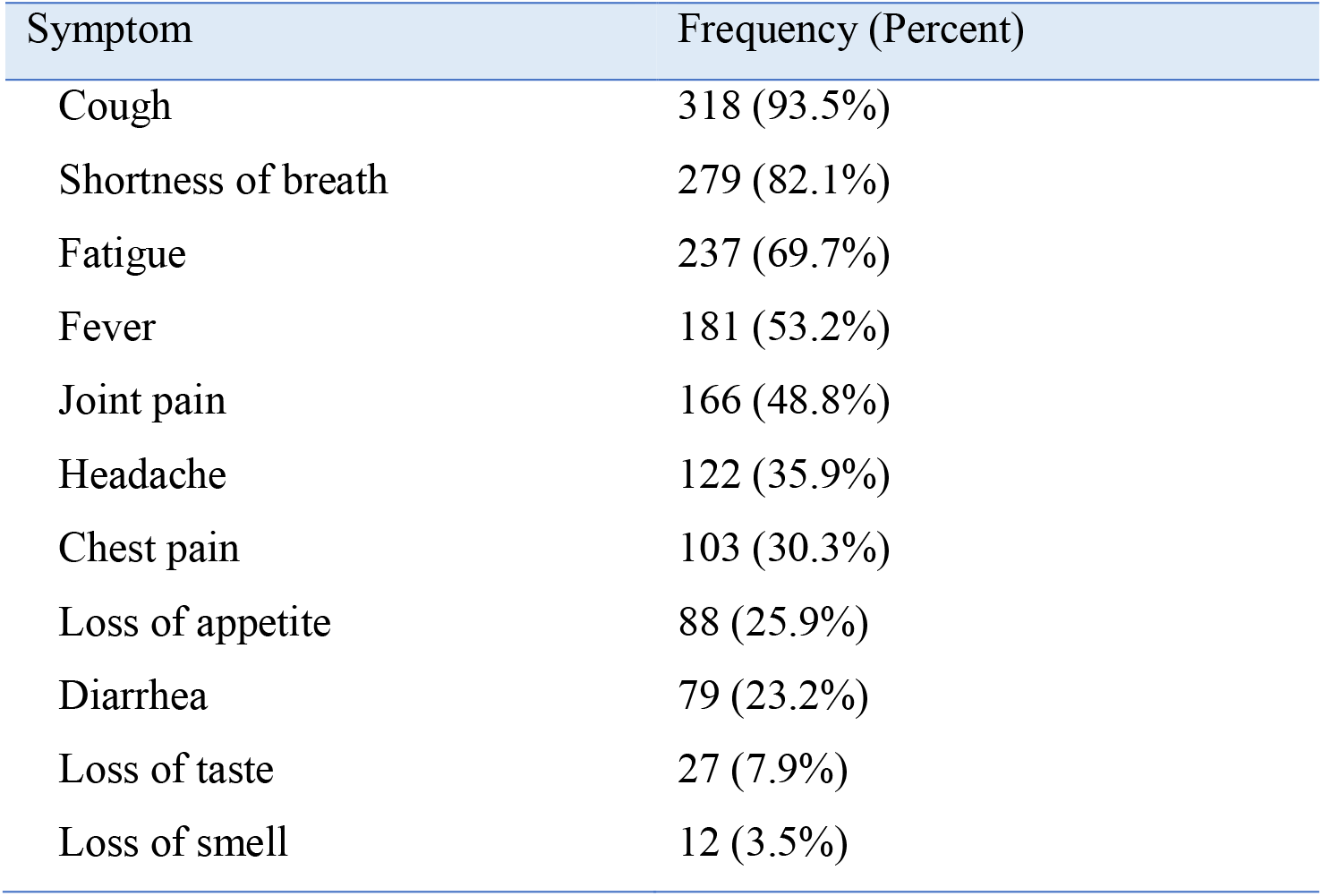
Presenting symptoms of patients who were admitted at Millennium COVID-19 care and treatment center with the diagnosis of severe COVID-19 pneumonia (n=340).

In the complete blood count (CBC) parameter of the patients, there were two outlier results recorded on patients with chronic lymphocytic leukemia (CLL), who had the white blood cell (WBC) count of 350,000 cells/ μL and 49,000 cells/ μL. The mean WBC count after excluding these two results was 9,148 (±4,044 SD) cells/ μL. The patients had a low mean lymphocyte percentage (10.2%). The mean hemoglobin was 14.6 (±1.7 SD) gm/dl and the mean platelet count was 285,840 (±116,588 SD) cells/ μL.

### Current status of study participants

Most (67.8%) of study participants visited a health facility at least once after their discharge from Millennium COVID-19 care and treatment center. The most common reason (59%) for the health facility visit was for follow-up of chronic disease and 15.7% of the reasons for hospital visit after discharge were not feeling well.

More than a third (38.1%) of the patients reported the persistence of at least one symptom after hospital discharge. The most common symptoms that started during the acute COVID-19 infection and continued till the time of the interview in descending order were; fatigue (27.5%), Cough (15.3%), joint pain (14.1%), headache (11.9%), and shortness of breath (11.3%). Symptoms that were less commonly found were diarrhea (2.5%) and loss of appetite (2.5%).

**Table 3:** Symptoms that have persisted after hospital discharge among patients who were admitted at Millennium COVID-19 care and treatment center with the diagnosis of severe COVID-19 pneumonia (n=320).

| Symptom | Frequency (Percent) |
| --- | --- |
| Fatigue | 88 (27.5%) |
| Cough | 49 (15.3%) |
| Joint pain | 45 (14.1%) |
| Headache | 38 (11.9%) |
| Shortness of breath | 36 (11.3%) |
| Sleep disturbance | 35 (10.9%) |
| Chest pain | 18 (5.6%) |
| Loss of appetite | 18 (5.6%) |
| Loss of smell | 18 (5.6%) |
| Diarrhea | 8 (2.5%) |
| Loss of taste | 8 (2.5%) |

Close to one-third (29.4%) of the patients feel that their health condition has deteriorated after the COVID-19 infection. Forty-four (13.8%) of the patients claimed that, currently they are not able to do the daily activities they used to do before the infection which forced some of the patients to change and even quit their job. Currently, three (0.9%) and fifty-five (17.2%) of the patients smoke cigarette and drink alcohol respectively. Of those who drink alcohol, most (40%) drink twice per month, followed by once per week (18.2%). Seven patients (12.7%) drink alcohol daily. Only 28.4% of the patients received at least one dose of vaccination.

### Factors associated with PACS

Patient age, sex, presence of comorbidity, alcohol use, baseline hemoglobin level, initial presentation with fatigue or loss of appetite, and length of hospital stay were found to be associated with the development of PACS in the patients at a significance level of P <0.05. However, cigarette smoking, current vaccination status, mean day of presentation after symptom onset, and maximum amount of oxygen used during the hospital stay were not found to influence the development of post-acute COVID-19 syndrome.

**Table 4:** Cross-tabulation of sociodemographic, clinical, and behavioral patterns with PACS among patients who were admitted at Millennium COVID-19 care and treatment center with the diagnosis of severe COVID-19 pneumonia (n=320).

| Characteristics | Has Symptom | Has no symptom | P Value |
| --- | --- | --- | --- |
| Mean Age (years) | 58.7 | 49.8 | < 0.01 |
| Sex |  |  | 0.01 |
| Male | 74 (33.6%) | 146 (66.4%) |  |
| Female | 48 (48%) | 52 (52%) |  |
| Comorbidity |  |  | < 0.01 |
| Yes | 91 (48.4%) | 97 (51.6%) |  |
| No | 31 (23.5%) | 101 (76.5%) |  |
| Specific Comorbidity |  |  |  |
| Hypertension | 56 (53.3%) | 49 (46.7%) | <0.01 |
| Diabetes | 48 (41.1%) | 68 (58.6%) | 0.36 |
| Dyslipidemia | 8 (66.7%) | 4 (33.3%) | 0.03 |
| Chronic kidney disease | 1 (100%) | 0 (0%) | 0.20 |
| Chronic liver disease | 3 (75%) | 1 (25%) | 0.12 |
| Chronic heart disease | 14 (87.5%) | 2 (12.5%) | <0.01 |
| Cancer | 4 (100%) | 0 (0%) | 0.01 |
| HIV | 6 (60%) | 4 (40%) | 0.14 |
| Stroke | 2 (66.7%) | 1 (33.3%) | 0.30 |
| Asthma or COPD | 8 (50%) | 8 (50%) | 0.31 |
| Cigarette smoking | 0 (0%) | 3 (100%) | 0.17 |
| Alcohol use | 28 (50.9%) | 27 (49.1%) | 0.03 |
| Mean length of hospital stay (days) | 14.0 | 11.2 | < 0.01 |
| Symptom duration before admission | 8.5 | 8.3 | 0.80 |
| Mean maximum amount of oxygen | 6.2 | 5.6 | 0.15 |
| Mean baseline hemoglobin | 14.3 | 14.8 | <0.01 |
| Current vaccination status |  |  | 0.34 |
| Vaccinated at least once | 31 (34.1%) | 60 (65.9%) |  |
| Not vaccinated | 91 (39.7%) | 138 (60.3%) |  |
| Fatigue at presentation | 93 (41.9%) | 129 (58.1%) | 0.03 |
| Loss of appetite at presentation | 24 (28.6%) | 60 (71.4%) | 0.03 |

By using variables that have a p-value of <0.25 in the bivariate analysis, binary logistic regression was done after the model fitness test. Factors that were independently associated with the development of PACS were older age (AOR 1.04, 95% CI 1.02 – 1.07), female sex (AOR 1.82, 95% CI 1.00 – 3.29), presence of comorbidity (AOR 2.38, 95% CI 1.35 – 4.19), alcohol use (AOR 3.05, 95% CI 1.49 – 6.26), fatigue at presentation (AOR 2.18, 95% CI 1.21 – 3.95), and longer hospital stay (AOR 1.06, 95% CI 1.02 – 1.10). Higher hemoglobin level was found to decrease the risk of subsequent PACS (AOR 0.84, 95% CI 0.71 – 0.99).

**Table 5:** Crude and adjusted odds ratio of factors that has a significant association with PACS among patients who were admitted at Millennium COVID-19 care and treatment center with the diagnosis of severe COVID-19 pneumonia (n=320).

| Characteristics | COR (95% CI) | P-Value | AOR (95% CI) | P-Value |
| --- | --- | --- | --- | --- |
| Age in years | 1.05 (1.03 - 1.08) | $<0.01$ | 1.04 (1.02 - 1.07) | $<0.01$ |
| Female sex | 1.82 (1.12 - 2.95) | 0.01 | 1.82 (1.00 - 3.29) | 0.04 |
| Presence of comorbidity | 3.05 (1.86 - 5.01) | $<0.01$ | 2.38 (1.35 - 4.19) | $<0.01$ |
| Drinking alcohol | 1.88 (1.05 - 3.38) | 0.03 | 3.05 (1.49 - 6.26) | $<0.01$ |
| Length of hospital stay | 1.06 (1.03 - 1.10) | $<0.01$ | 1.06 (1.02 - 1.10) | 0.01 |
| Hemoglobin level | 0.82 (0.71 - 0.95) | $<0.01$ | 0.84 (0.71 - 0.99) | 0.04 |
| Fatigue at presentation | 1.71 (1.03 – 2.85) | 0.03 | 2.18 (1.21 -3.95) | $<0.01$ |
| Loss of appetite | 0.56 (0.32 – 0.96) | 0.03 | 0.59 (0.31 – 1.09) | 0.09 |

## Discussion

As to the Authors’ knowledge, this is the first study that assessed the health consequences of COVID-19 at two-year follow-up in patients who had severe COVID-19 pneumonia. 340 patients who were admitted to Millennium COVID-19 care and treatment center with the diagnosis of severe COVID-19 pneumonia were included in the study. The majority (68.5%) were males and the mean age was 53.9 (±13.3 SD) years. More than half of the patients (60%) has one or more comorbidity. The most common symptom at presentation was cough (93.5%), followed by shortness of breath (82.1%) and fatigue (69.7%). The mean duration of hospital admission was 12.3 (±6.5 SD) days. More than a third (38.1%) of the patients reported the persistence of at least one symptom after hospital discharge. The most common symptoms were fatigue (27.5%) and Cough (15.3%). Older age, female sex, presence of comorbidity, alcohol use, low baseline hemoglobin level, fatigue at presentation, and prolonged hospital stay were found to increase the odds of developing PACS.

The patients in this study were younger and males were more represented when compared to other similar studies. The mean age (53.9 years) at presentation was younger by six years in this study when compared to the cohort study conducted in Italy (19). This is probably due to the demographic background of Ethiopia, where the proportion of the elderly population is lower than that in the Western world. The male-to-female ratio was 1.4:1 in the Bangladesh study, but the ratio is significantly higher (2.1:1) in this study (9).

The death rate after hospital discharge in this study was lower (5.8%) compared to a study done in Spain on patients who were admitted to Hospital for COVID-19. The study done in Spain found that 12.8% of the patients died within a mean follow-up period of one year (22).

The study participants in this study have a significantly higher level of comorbidity (60%) when compared to the cross-sectional study in Egypt on 430 patients found that 26.5% patients reported that they have a chronic illness, and the Norwegian prospective cohort study, in which, 44% had comorbidities (14, 23). This significant difference in comorbidity is observed mainly because the other studies were done on all COVID-19 patients, and this study was done specifically on patients with severe COVID-19 pneumonia. Diabetes (35.6%), hypertension (34.1%), and chronic heart disease (6.5%) were the most common comorbidities in this study which is similar to the cohort studies done in China and England (16, 24).

The prevalence of PACS in this study was 38.1%, which is lower than the finding in most of the studies, which were in the range of 46% in Bangladesh to 87.4% in Italy. (9, 25) This may be because, in those studies, the maximum follow-up period was one year, but this study was conducted after a mean period of 25.6 months after hospital discharge and symptoms might have improved over time. The causes underlying these persistent symptoms following COVID-19 are not entirely understood. In addition to the direct effects of SARS-CoV-2, the immunological response to the virus is thought to have a role in the development of these long-term symptoms, presumably by supporting a continuing hyper-inflammatory process (26).

Fatigue is the most common (27.5%) symptom of PACS in this study, which is in concordance with the findings of most other similar studies (13, 14, 25). Although the exact cause and pathogenesis of fatigue following COVID-19 is unknown, previous data from severe acute respiratory syndrome (SARS) suggests that lung diffusion capacity impairment, some extra-pulmonary causes, such as viral-induced myositis at initial presentation, cytokine disturbance, muscle wasting, and deconditioning, or corticosteroids myopathy, or a combination of these factors, may have contributed to the condition (27).

The second most common symptom in this study was cough (15.3%). This finding mirrors the findings of previous similar studies (9, 18, 28). The mechanisms of cough after COVID-19 are multifactorial, including parenchymal sequelae and activation of the vagal sensory nerves, which leads to a cough hypersensitivity state (29).

In this study, 48% of female patients reported the presence of symptoms at the time of the interview. Female sex (AOR = 1.82, 95% CI 1.00 - 3.29, P=0.04) was found to increase the risk of developing PACS. This findings are similar to other studies (11, 12, 30). Various underlying processes explaining why females experience post-COVID symptoms to a larger extent than males are now being studied in the literature. Male and female biological differences in the expression of angiotensin-converting enzyme-2 (ACE2) and transmembrane protease serine 2 (TMPRSS2) receptors, as well as immunological differences, such as lower production of pro-inflammatory interleukin-6 (IL-6) after viral infection in females, could explain the higher development of post-COVID symptoms (31).

Older age (AOR = 1.04, 95% CI 1.02 – 1.07, P: <0.01) was found to be a statistically significant predictor for the development of PACS. This is a similar finding to a study done in France where older age increased the risk of subsequent PACS (AOR=1.49, 95% CI 1.05–2.17) (15).

Unlike the Bangladesh cohort study which showed patients with fever, cough, respiratory distress, and lethargy as the presenting features were more susceptible to develop PACS compared to other presenting features and the Indian study which showed diarrhea at presentation to be associated with PACS, the only presenting feature that was found to be significant in this study was fatigue (AOR = 2.18, 95% CI 1.21 - 3.95, P: <0.01) (9, 32).

In this study, prolonged hospital stay was found to significantly increase the risk of PACS (AOR = 1.06, 95% CI 1.02 - 1.10, P=0.01). A similar finding was observed in a study conducted in Spain which revealed that the number of days at the hospital was significantly associated with an increased risk of PACS (33).

Although the number of days between symptom onset and admission in the Indian study and the amount of oxygen requirement in the Egyptian study were found to determine PACS, this study found no association between those factors and PACS (17, 32).

Another finding in our study was that COVID-19 vaccination was not found to be protective from PACS. This was also shown in previous studies (9).

The studies on the effect of smoking as a risk factor for developing post-acute COVID-19 syndrome showed conflicting results. A study conducted in Egypt showed that there is no significant association between cigarette smoking and post-acute COVID-19 syndrome (18), while studies conducted in China and Spain study found a strong association between current active smoking and post-acute COVID-19 syndrome (10, 16). This study found no significant association between current cigarette smoking and PACS.

The effect of alcohol intake on the development of PACS is found to be significant (AOR = 3.05 [1.49 – 6.26], P: <0.01) in this study, which is a similar finding to the Mediterranean cohort study and the Bangladesh study (9, 13).

## Conclusion

The prevalence of PACS syndrome among severe COVID-19 patients who were discharged alive from Millennium COVID-19 Care and treatment center between June 12, 2020, and November 15, 2021, after a mean period since discharge of 25.6 months, was found to be 38.1%. Fatigue (27.5%) and cough (15.3%) were the most prevalent symptoms. Older age, female sex, presence of comorbidity, alcohol use, low baseline hemoglobin level, fatigue at presentation, and prolonged hospital stay were found to increase the odds of developing PACS.

## Data Availability

All data produced in the present study are available upon reasonable request to the authors.

## Notes

### Competing Interest Statement

The authors have declared no competing interest.

### Funding Statement

The study did not receive any funding.

### Author Declarations

IRB of St. Paul's hospital millennium medical college gave ethical approval for this work.

## References

1. Who’s Response To Covid-19. 2021;(January).

2. WHO Coronavirus (COVID-19) Dashboard. Available at: https://covid19.who.int.

3. Kabi A, Mohanty A, Mohanty AP, Kumar S. Post COVID-19 Syndrome: A Literature Review. JAdv Med Med Res. 2020;32(24):289–95.

4. Nalbandian A, Sehgal K, Gupta A, Madhavan M V., McGroder C, Stevens JS, et al. Post-acuteCOVID-19 syndrome. Nat Med. 2021;27(4):601–15.

5. Oronsky B, Larson C, Hammond TC, Oronsky A, Kesari S, Lybeck M, et al. A Review of Persistent Post-COVID Syndrome (PPCS). Clin Rev Allergy Immunol. 2021;

6. Shah W, Hillman T, Playford ED, Hishmeh L. Managing the long term effects of covid-19: Summary of NICE, SIGN, and RCGP rapid guideline. BMJ. 2021; 372:103.

7. Fernández-De-las-peñas C, Palacios-Ceña D, Gómez-Mayordomo V, Cuadrado ML, FlorencioLL. Defining post-covid symptoms (Post-acute covid, long covid, persistent post-covid): An integrative classification. Int J Environ Res Public Health. 2021; 18(5):1–9.

8. UK NHS. National Guidance for post-COVID syndrome assessment clinics. Version 2.2021;(April):1–44.

9. Mahmud R, Rahman MM, Rassel MA, Monayem FB, Sayeed SKJB, Islam MS, et al. Post-COVID-19 syndrome among symptomatic COVID-19 patients: A prospective cohort study in atertiary care center of Bangladesh. PLoS One. 2021;16(4 April):1–13.

10. Michele Davide Maria Lombardo and others, Long-Term Coronavirus Disease 2019 Complications in Inpatients and Outpatients: A One-Year Follow-up Cohort Study, Open Forum Infectious Diseases, Volume 8, Issue 8, August 2021, ofab384, https://doi.org/10.1093/ofid/ofab384

11. Sultana S, Islam MT, Salwa M, et al. Duration and Risk Factors of Post-COVID Symptoms Following Recovery Among the Medical Doctors in Bangladesh. Cureus. 2021;13(5):e15351. Published 2021 May 31. doi:10.7759/cureus.15351

12. Louis Jacob, Ai Koyanagi, Lee Smith, Christian Tanislav, Marcel Konrad, Susanne van der Beck, Karel Kostev, et al. Prevalence of, and factors associated with, long-term COVID-19 sick leave in working-age patients followed in general practices in Germany, International Journal of Infectious Diseases, Volume 109, 2021, Pages 203–208, https://doi.org/10.1016/j.ijid.2021.06.063

13. Moreno-pérez O, Merino E, Leon-ramirez J, Andres M, Manuel J, Arenas-jiménez J, et al. Post-acute COVID-19 syndrome. Incidence and risk factors: A Mediterranean cohort study. J Infect. 2021;82(January):373–8.

14. Blomberg B, Mohn KGI, Brokstad KA, Zhou F, Linchausen DW, Hansen BA, et al. Long COVIDin a prospective cohort of home-isolated patients. Nat Med. 2021;

15. Carvalho-schneider C, Laurent E, Lemaignen A, Laribi S, Flament T, Beau E, et al. Follow-up of adults with noncritical COVID-19 two months after symptom onset. 2020;(January).

16. Huang C, Huang L, Wang Y, Li X, Ren L, Gu X, et al. 6-month consequences of COVID-19 in patients discharged from hospital: a cohort study. Lancet [Internet]. 2021;397(10270):220–32. Available from: http://dx.doi.org/10.1016/S0140-6736(20)32656-8

17. Galal I, Hussein AARM, Amin MT, et al. Determinants of persistent post-COVID-19 symptoms: value of a novel COVID-19 symptom score. The Egyptian Journal of Bronchology. 2021;15(1):10. doi:10.1186/s43168-020-00049-4

18. Galal I, Hussein AARM, Amin MT, Saad MM, Zayan HEE, Abdelsayed MZ, et al. Determinants of persistent post-COVID-19 symptoms: value of a novel COVID-19 symptom score. Egypt J Bronchol. 2021;15(1).

19. Michele Davide Maria Lombardo and others, Long-Term Coronavirus Disease 2019 Complications in Inpatients and Outpatients: A One-Year Follow-up Cohort Study, Open Forum Infectious Diseases, Volume 8, Issue 8, August 2021, ofab384, https://doi.org/10.1093/ofid/ofab384

20. Gudina EK, Tesfaye M, Siraj D, Haileamilak A, Yilma D. COVID-19 in Ethiopia in the first 180days: Lessons learned and the way forward. Ethiop J Heal Dev. 2020;34(4):301–6.

21. Global COVID-19 Clinical Platform Case Report Form (CRF) for Post COVID condition (Post COVID-19 CRF) [Internet]. 2021 Feb [cited 2021 Dec 13]. Available from: https://www.who.int/publications/i/item/global-covid-19-clinical-platform-case-report-form-(crf)-for-post-covid-conditions-(post-covid-19-crf-).

22. Maestre-Muñiz MM, Arias Á, Mata-Vázquez E, Martín-Toledano M, López-Larramona G, Ruiz-Chicote AM, et al. Long-Term Outcomes of Patients with Coronavirus Disease 2019 at One Year after Hospital Discharge. Journal of Clinical Medicine 2021;10(13):2945. Available from: http://dx.doi.org/10.3390/jcm10132945

23. Kamal M, Abo Omirah M, Hussein A, Saeed H. Assessment and characterisation of post-COVID-19 manifestations. Int J Clin Pract. 2021;75(3):0–2.

24. Docherty AB, Harrison EM, Green CA, Hardwick HE, Pius R, Norman L, et al. Features of 20133 UK patients in hospital with covid-19 using the ISARIC WHO Clinical Characterisation Protocol: Prospective observational cohort study. BMJ. 2020;369:1–2.

25. Carfì A, Bernabei R, Landi F. Persistent symptoms in patients after acute COVID-19. JAMA - JAm Med Assoc. 2020;324(6):603–5.

26. Mohamad Salim Alkodaymi, Osama Ali Omrani, et al. Prevalence of post-acute COVID-19 syndrome symptoms at different follow-up periods: a systematic review and meta-analysis, Clinical Microbiology and Infection, Volume 28, Issue 5, 2022, Pages 657-666,ISSN 1198-743X, https://doi.org/10.1016/j.cmi.2022.01.014.

27. Lixue Huang, Qun Yao, Xiaoying Gu, et al. 1-year outcomes in hospital survivors with COVID-19: a longitudinal cohort study, The Lancet, Volume 398, Issue 10302, 2021, Pages 747-758, ISSN 0140-6736, https://doi.org/10.1016/S0140-6736(21)01755-4.

28. Tenforde MW, Kim SS, Lindsell CJ, Billig Rose E, Shapiro NI, Files DC, et al. Symptom Duration and Risk Factors for Delayed Return to Usual Health Among[1] M. W. Tenforde et al.,”Symptom Duration and Risk Factors for Delayed Return to Usual Health Among Outpatients with COVID-19 in a Multistate Health Care Systems Network — United. MMWR Morb MortalWkly Rep. 2020;69(30):993–8.

29. Montani D, Savale L, Noel N, et al. Post-acute COVID-19 syndrome. Eur Respir Rev 2022; 31: 210185 [DOI: 10.1183/16000617.0185-2021].

30. Caroline E. Gebhard, Claudia Sütsch, Susan Bengs, Manja Deforth, Karl Philipp Buehler, et al. Sex- and Gender-specific Risk Factors of Post-COVID-19 Syndrome: A Population-based Cohort Study in Switzerland. medRxiv 202106.30; doi:https://doi.org/10.1101/2021.06.30.21259757

31. Fernández-de-las-Peñas, César et al. “Female Sex Is a Risk Factor Associated with Long-Term Post-COVID Related-Symptoms but Not with COVID-19 Symptoms: The LONG-COVID-EXP-CM Multicenter Study.” Journal of Clinical Medicine 11 (2022)

32. Neha Chopra, Mohit Chowdhury, Anupam K Singh, Khan MA, Arvind Kumar, et al., Clinical predictors of long COVID-19 and phenotypes of mild COVID-19 at a tertiary care centre in India, Drug Discoveries &Therapeutics, 2021, Volume 15, Issue 3, Pages 156–161, Released July 06, 2021, https://doi.org/10.5582/ddt.2021.01014

33. Fernández-de-Las-Peñas C, Palacios-Ceña D, Gómez-Mayordomo V, et al. Long-term post-COVID symptoms and associated risk factors in previously hospitalized patients: A multicenter study. J Infect. 2021;83(2):237–279. doi:10.1016/j.jinf.2021.04.036

